# The effect of the COVID-19 lockdown on mental health care use in South Africa: an interrupted time series analysis

**DOI:** 10.1101/2022.04.07.22273561

**Authors:** Anja Wettstein, Mpho Tlali, John A Joska, Morna Cornell, Veronika W Skrivankova, Soraya Seedat, Johannes P Mouton, Leigh L van den Heuvel, Nicola Maxwell, Mary-Ann Davies, Gary Maartens, Matthias Egger, Andreas D Haas

**Affiliations:** Institute of Social and Preventive Medicine, University of Bern, Bern, Switzerland; Graduate School for Health Sciences, University of Bern, Switzerland; Centre for Infectious Disease Epidemiology and Research, University of Cape Town, Cape Town, South Africa; HIV Mental Health Research Unit, Department of Psychiatry and Mental Health, Neuroscience Institute, Cape Town, South Africa; Department of Psychiatry, Faculty of Medicine and Health Sciences, Stellenbosch University, Cape Town, South Africa; Division of Clinical Pharmacology, Department of Medicine, University of Cape Town, Cape Town, South Africa; Population Health Sciences, Bristol Medical School, University of Bristol, Bristol, UK

**Author notes:** Corresponding author: Andreas Haas, PhD, Institute of Social and Preventive Medicine (ISPM), University of Bern, Mittelstrasse 43 CH-3012 Bern.

## Abstract

**Aims:** In March 2020, South Africa introduced a lockdown in response to the COVID-19 pandemic, entailing the suspension of all non-essential activities and a complete ban of tobacco and alcohol sales. We studied the effect of the lockdown on mental health care utilisation rates in private-sector care in South Africa.

**Methods:** We did an interrupted time series analysis using insurance claims from January 1, 2017, to June 1, 2020 of beneficiaries 18 years or older from a large private sector medical aid scheme. We calculated weekly outpatient consultation and hospital admission rates for organic mental disorders, substance use disorders, serious mental disorders, depression, anxiety, other mental disorders, any mental disorder, and alcohol withdrawal syndrome. We calculated adjusted odds ratios (OR) for the effect of the lockdown on weekly outpatient consultation and hospital admission rates and the weekly change in rates during the lockdown until June 1, 2020.

**Results:** 710,367 persons were followed up for a median of 153 weeks. Hospital admission rates (OR 0.38; 95% CI 0.33–0.44) and outpatient consultation rates (OR 0.74; 95% CI 0.63–0.87) for any mental disorder decreased substantially after the lockdown and did not recover to pre-lockdown levels until June 1, 2020. Health care utilisation rates for alcohol withdrawal syndrome doubled after the introduction of the lockdown, but the statistical uncertainty around the estimates was large (OR 2.24; 95% CI 0.69-7.24).

**Conclusions:** Reduced mental health care contact rates during the COVID-19 lockdown likely reflect a substantial unmet need for mental health services with potential long-term consequences for mental health patients and their families. Steps to ensure access and continuity of mental health services during future lockdowns should be considered.

## Introduction

On March 5, 2020, South Africa reported its first COVID-19 case, which was followed by the detection of clusters of cases and high rates of community transmission (Moonasar *et al*. 2021). In response to the pandemic, South Africa introduced a stringent set of restrictions, called (level 5) “lockdown” on March 27, 2020, entailing the suspension of all non-essential activities and a complete ban of tobacco and alcohol sales. On May 1, 2020, restrictions were eased to level 4, allowing people to buy more than essential goods, have food delivered, and exercise outside for a brief period. With the move to level 3 on June 1, 2020, limited alcohol sales were allowed, and more businesses could open, but the beauty and tourism sectors remained closed (Greyling *et al*. 2021).

The COVID-19 pandemic and lockdowns negatively impact the mental health and well-being of the general population (Brooks *et al*. 2020; Liao *et al*. 2021; Santomauro *et al*. 2021; Winkler *et al*. 2020; Xiong *et al*. 2020). A systematic review reported high rates of symptoms of anxiety, depression, post-traumatic stress disorder, and psychological distress during the COVID-19 pandemic (Xiong *et al*. 2020). Fear and uncertainty associated with the COVID-19 pandemic, and the drastic implications of the response to the pandemic on people’s life and the economy, including social isolation, loneliness, confinement, physical inactivity, frustration, boredom, limited access to basic supplies and services, loss of jobs and financial worries, exacerbate the risk of incident of mental health disorders and the severity of existing mental health conditions (Moreno *et al*. 2020).

There is increasing evidence suggesting that the response to the COVID-19 pandemic led to a disruption of health services. Several studies, mainly from Europe, North America, and Asia, reported a substantial decrease in the rates of emergency department visits (Jeffery *et al*. 2020; Wongtanasarasin *et al*. 2021) and hospital admissions for acute medical conditions including cardiovascular diseases (Esenwa *et al*. 2020; Pelletier *et al*. 2021) and mental health problems (Boldrini *et al*. 2021; Gale *et al*. 2021; Gómez-Ramiro *et al*. 2021; McDowell *et al*. 2021; Wyatt *et al*. 2021) following the introduction of COVID-19 lockdowns. The effect of COVID-19 related lockdowns on outpatient care is less well researched. Studies from high-income countries report reduced outpatient care contacts for physical and mental health conditions during the COVID-19 pandemic (Mansfield *et al*. 2021; Seo *et al*. 2021; Williams *et al*. 2020). Less is known from the low- and middle-income country context (Kola *et al*. 2021). A study from South Africa found a sharp decline in HIV testing and antiretroviral therapy initiation rates but no decline in antiretroviral therapy collection visits in primary care HIV clinics after the lockdown (Dorward *et al*. 2021). The effect of the lockdown on inpatient and outpatient mental health care utilisation in African countries is unclear.

We aimed to quantify the impact of the COVID-19 lockdown (levels 5 and 4) on mental health care utilisation in private sector care in South Africa. We assessed the effect of lockdown measures on weekly hospital admission and outpatient consultation rates for selected mental disorders. In addition, we tested the hypothesis that the ban on alcohol sales led to an increased rate of hospital admissions and outpatient consultations for alcohol withdrawal syndrome.

## Methods

### Study design

We conducted an interrupted time series analysis on the effects of the level 5 and 4 COVID-19 lockdown on mental health care use in South Africa’s private health sector using outpatient and hospital claim data with corresponding ICD10 diagnoses from a large private sector medical scheme. We analysed data from January 1, 2017, to June 28, 2020. We adopted the study design from a previous study evaluating the effect of COVID-19 measures on health care use in the UK.(Mansfield *et al*. 2021) The Human Research Ethics Committee of the University of Cape Town and the Cantonal Ethics Committee of the Canton of Bern granted permission to analyse the data.

### Study population

We followed beneficiaries of one of South Africa’s largest open medical schemes that insured over 700,000 individuals as of 2019.(Council for Medical Schemes 2020) It has a young membership base with an average age of about 33.(Council for Medical Schemes 2020) We included beneficiaries aged 18 years or older who had an active health care plan between January 1, 2017, and June 28, 2020.Individuals with missing information on sex or age were excluded. Follow-up ended at termination of the insurance contract, the date of death, or the end date of the study period.

### Exposures, outcomes, and stratifying variables

The exposure of interest was the introduction of the national lockdown in South Africa on March 27, 2020. We defined the start of the lockdown as the beginning of week 14 (March 30, 2020).

Outcomes were the proportion of beneficiaries (1) admitted to a hospital, (2) consulting outpatient care, or (3) receiving any mental health care (either being admitted to a hospital or consulting outpatient care) for selected mental disorders. The South African Health Profession Council embraced telemedicine to overcome shortages in health care delivery and to protect health care staff on March 26, 2020. With the amendments of April 3, 2020, telemedicine could also be used for first-time consultations and allowed for reimbursement for telemedical services through the insurance system.(Kwinda 2020) Our definition of outpatient care consultations, therefore, included both, in-person and telemedical consultations.

We identified mental disorders based on ICD10 diagnoses from outpatient and hospital claims: organic mental disorders (ICD codes F00-09), substance use disorders (F10-F19), serious mental disorders such as Schizophrenia spectrum disorders, psychotic, delusional, or bipolar disorders (F20-F29, and F31), depressive disorders (F32, F34.1, and F54) anxiety and related disorders (F40-F48), other mental disorders like a single manic episode, persistent mood affective disorders, eating disorders, sleep disorders, or unspecified mental disorders (F30, F34.0, F34.8, F34.9, F50-F53, and F55-99), and alcohol withdrawal syndrome (F10.3, and F10.4). Finally, we defined any mental disorder as being diagnosed with any ICD10 F00-F99 diagnosis.

### Statistical analysis

We described the sociodemographic characteristics of the study population under follow-up on January 1 of each year using summary statistics. We calculated and plotted weekly mental health care utilisation rates defined as the percentage of beneficiaries receiving care for a defined condition in each week between January 1, 2017, and June 28, 2020.

We conducted interrupted time-series analyses to assess changes in weekly mental health care utilisation rates during the level 5 and 4 COVID-19 lockdowns. In these analyses, we did not use data from the last four weeks before database closure (June 1, 2020 – June 28, 2020) to account for delays in reporting. In addition, we did not use data from weeks 12 to week 13 (March 15 – March 29, 2020) to account for the anticipatory behaviour of beneficiaries following the announcement of the National State of Disaster in South Africa on March 15, 2020. The interrupted time series analysis assumes that under the counterfactual scenario, the pre-lockdown time series continues during the lockdown and compares the extrapolated pre-lockdown time series to the observed post-lockdown time series. We modelled weekly health care utilisation rates using binomial generalized linear regression models with logit link and robust standard errors (Papke & Wooldridge 1996). Models included a linear effect of time and an indicator variable for calendar months to account for long-term trends and seasonal variation in mental health care use. In addition, models included a binary indicator for the lockdown to measure the immediate change in health care use following the implementation of the lockdown and an interaction term between time and the binary indicator to measure the slope change in health care use during the lockdown (Mansfield *et al*. 2021). Results are presented as odds ratios (ORs) for the immediate effect of the lockdown on mental health care use and for the weekly change in the utilisation during the lockdown period (Mansfield *et al*. 2021). We stratified interrupted time series analysis of the change in mental health care use for any mental disorders by sex. In sensitivity analysis, we implemented the model used by Mansfield and colleagues and compared results to our primary analysis (Mansfield *et al*. 2021). The Mansfield model uses conventional standard errors and adjusts for autocorrelation by including first-order lagged residuals (Mansfield *et al*. 2021). Finally, to validate our model, we performed the same interrupted time-series analysis (week 14-22) of mental health care utilisation for 2019, expecting no changes in this period. We implemented the Mansfield model in R version 4.1.1 (R Core Team 2021). All other analyses were done in Stata version 16 (StataCorp, College Station, TX, USA). Statistical code is available under https://github.com/AndreasDHaas/ECMHC.

## Results

Of 1,013,033 beneficiaries who had an active health care plan with the medical aid scheme during the study period, 710,367 were eligible for analysis. We excluded 296,155 children and adolescents aged 17 years or younger at the end of their follow-up and 6,511 beneficiaries with incomplete data on sex and age. The median follow-up time was 153 weeks (interquartile range [IQR] 57-178). At the beginning of 2017, 53% of the study population were women, and the median age was 43 years (IQR 32-56) (Table 1). The number of beneficiaries, and their age, sex, and population group distributions, remained relatively stable throughout the study period.

**Table 1:**
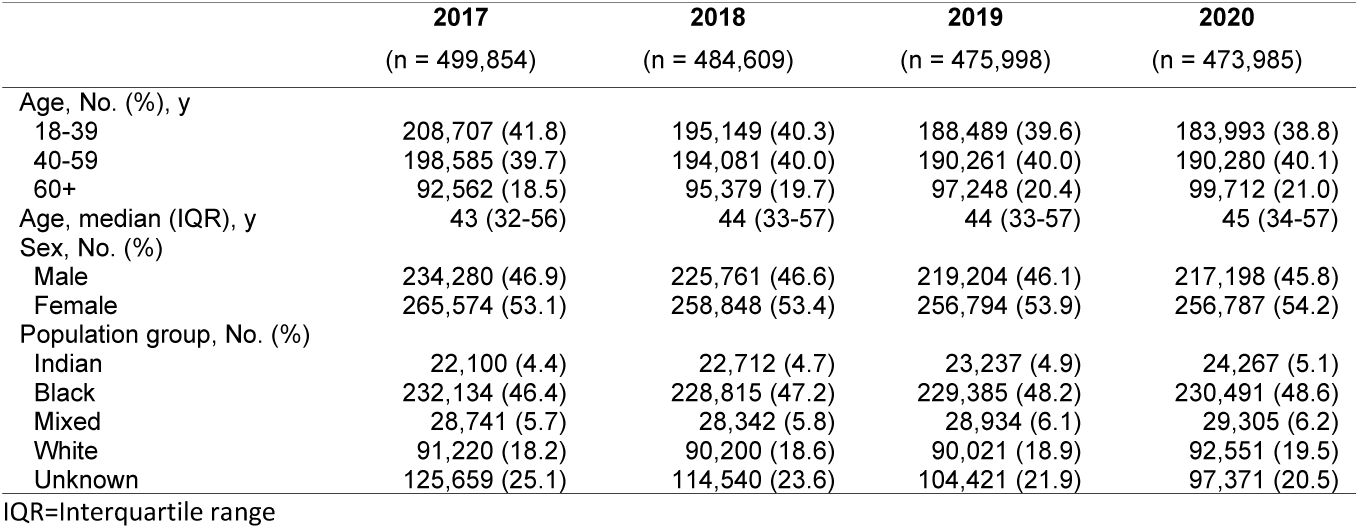
Characteristics of the study population at the beginning of each year, 2017-2020.

|  | 2017 | 2018 | 2019 | 2020 |
| --- | --- | --- | --- | --- |
|  | (n = 499,854) | (n = 484,609) | (n = 475,998) | (n = 473,985) |
| Age, No. (%), y |  |  |  |  |
| 18-39 | 208,707 (41.8) | 195,149 (40.3) | 188,489 (39.6) | 183,993 (38.8) |
| 40-59 | 198,585 (39.7) | 194,081 (40.0) | 190,261 (40.0) | 190,280 (40.1) |
| 60+ | 92,562 (18.5) | 95,379 (19.7) | 97,248 (20.4) | 99,712 (21.0) |
| Age, median (IQR), y | 43 (32-56) | 44 (33-57) | 44 (33-57) | 45 (34-57) |
| Sex, No. (%) |  |  |  |  |
| Male | 234,280 (46.9) | 225,761 (46.6) | 219,204 (46.1) | 217,198 (45.8) |
| Female | 265,574 (53.1) | 258,848 (53.4) | 256,794 (53.9) | 256,787 (54.2) |
| Population group, No. (%) |  |  |  |  |
| Indian | 22,100 (4.4) | 22,712 (4.7) | 23,237 (4.9) | 24,267 (5.1) |
| Black | 232,134 (46.4) | 228,815 (47.2) | 229,385 (48.2) | 230,491 (48.6) |
| Mixed | 28,741 (5.7) | 28,342 (5.8) | 28,934 (6.1) | 29,305 (6.2) |
| White | 91,220 (18.2) | 90,200 (18.6) | 90,021 (18.9) | 92,551 (19.5) |
| Unknown | 125,659 (25.1) | 114,540 (23.6) | 104,421 (21.9) | 97,371 (20.5) |

IQR=Interquartile range

Hospital admission rates for any mental disorder decreased substantially after the introduction of the lockdown (OR 0.38; 95% CI 0.33–0.44) and did not recover to pre-lockdown levels until June 1, 2020 (Figure 1). Except for alcohol withdrawal syndrome (OR 1.36; 95% CI 0.46-4.02) and other mental disorders (OR 0.36; 95% 0.12-1.10) admission rates for all mental disorders decreased significantly.

**Figure 1:**
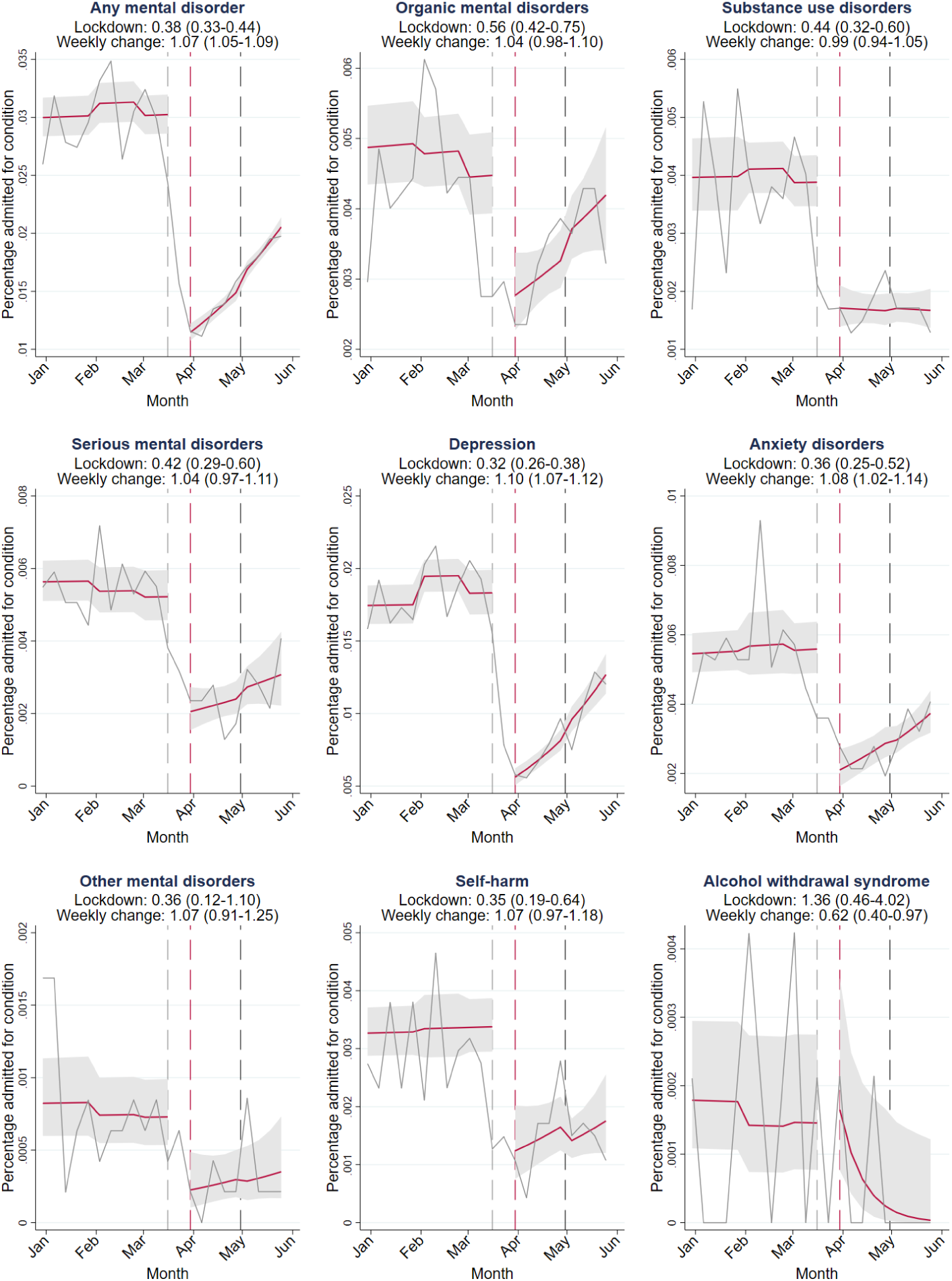
Interrupted time-series analysis for changes in hospital admissions during the lockdown. Solid grey lines represent percentages of the study population admitted for the condition in each week between January 1, 2020, and June 1, 2020. Solid red lines depict the estimated average percentage admitted per week with 95% confidence intervals (grey shaded areas). Area between the dashed grey and dashed red line: data not used to account for anticipatory behaviour. Dashed red line: first Monday during lockdown level 5 (March 30, 2020). Dashed black line: beginning of lockdown level 4 (April 30, 2020). Lockdown: odds ratios (OR) for the immediate effect of the lockdown on admission rates. Weekly change: OR for the weekly change in the odds of hospital admission during the lockdown (week 14-22 in 2020). 95% confidence intervals for ORs in parentheses.

Outpatient consultation rates for any mental disorder decreased after the lockdown was introduced (OR 0.74; 95% CI 0.63–0.87) and did not fully recover to pre-pandemic levels during the study period (Figure 2). There was no strong evidence of an effect of the lockdown on outpatient consultation rates for self-harm. Rates of outpatient consultations for alcohol withdrawal syndrome doubled after the introduction of the lockdown, but the statistical uncertainty around the estimates was large (OR 2.11; 95% CI 0.53-8.43). Outpatient consultation rates for most categories recovered after the initial drop, but rates for alcohol withdrawal syndrome declined during the lockdown.

**Figure 2:**
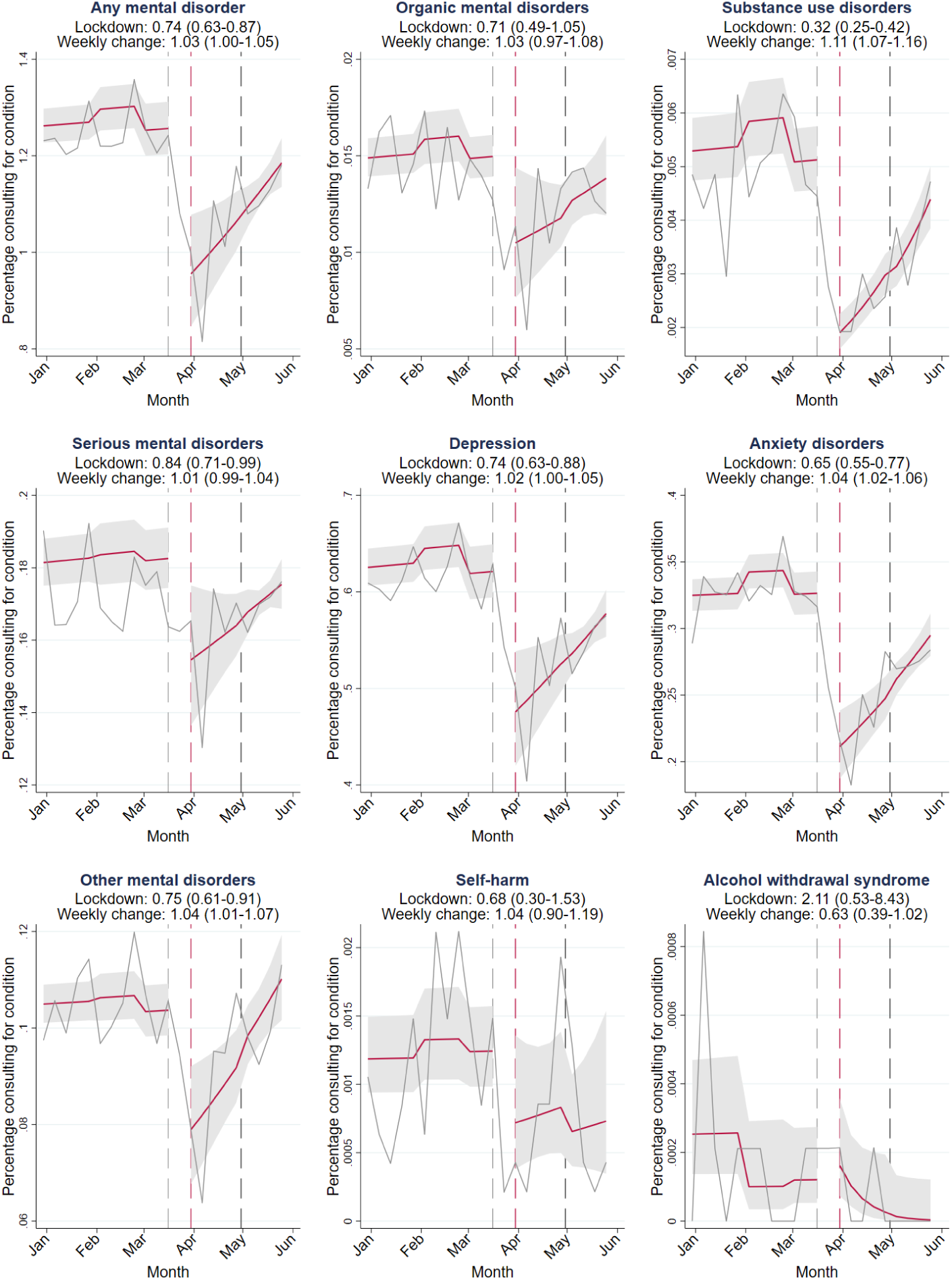
Interrupted time-series analysis for changes in outpatient consultations during the lockdown. Solid grey lines represent percentages of the study population consulting outpatient care for the condition in each week between January 1, 2020, and June 1, 2020. Red lines depict the estimated average percentage consulting outpatient care per week with 95% CIs (grey shaded areas). Area between the dashed grey and dashed red line: data not used to account for anticipatory behaviour. Dashed red line: first Monday during lockdown (March 30, 2020). Dashed black line: beginning of lockdown level 4 (April 30, 2020). Lockdown: odds ratios (OR) for the immediate effect of the lockdown on outpatient consultation rates. Weekly change: OR for the weekly change in the odds of outpatient consultation rates during the lockdown (week 14-22 in 2020). 95% confidence intervals for ORs in parentheses.

Results from the interrupted time series analysis of the overall mental health care utilisation rates including hospital admission and outpatient consultation rates are shown in Figure 3. Similar to outpatient consolation rates, overall mental health care utilisation rates decreased after the lockdown was introduced (OR 0.74; 95% CI 0.63–0.86) and did not fully recover to pre-pandemic levels during the study period. The combined rates of hospital admissions and outpatient care consultations for alcohol withdrawal syndrome doubled after the introduction of the lockdown, but the statistical uncertainty around the estimates of the combined rates remained large (OR 2.24; 95% CI 0.69-7.24).

**Figure 3:**
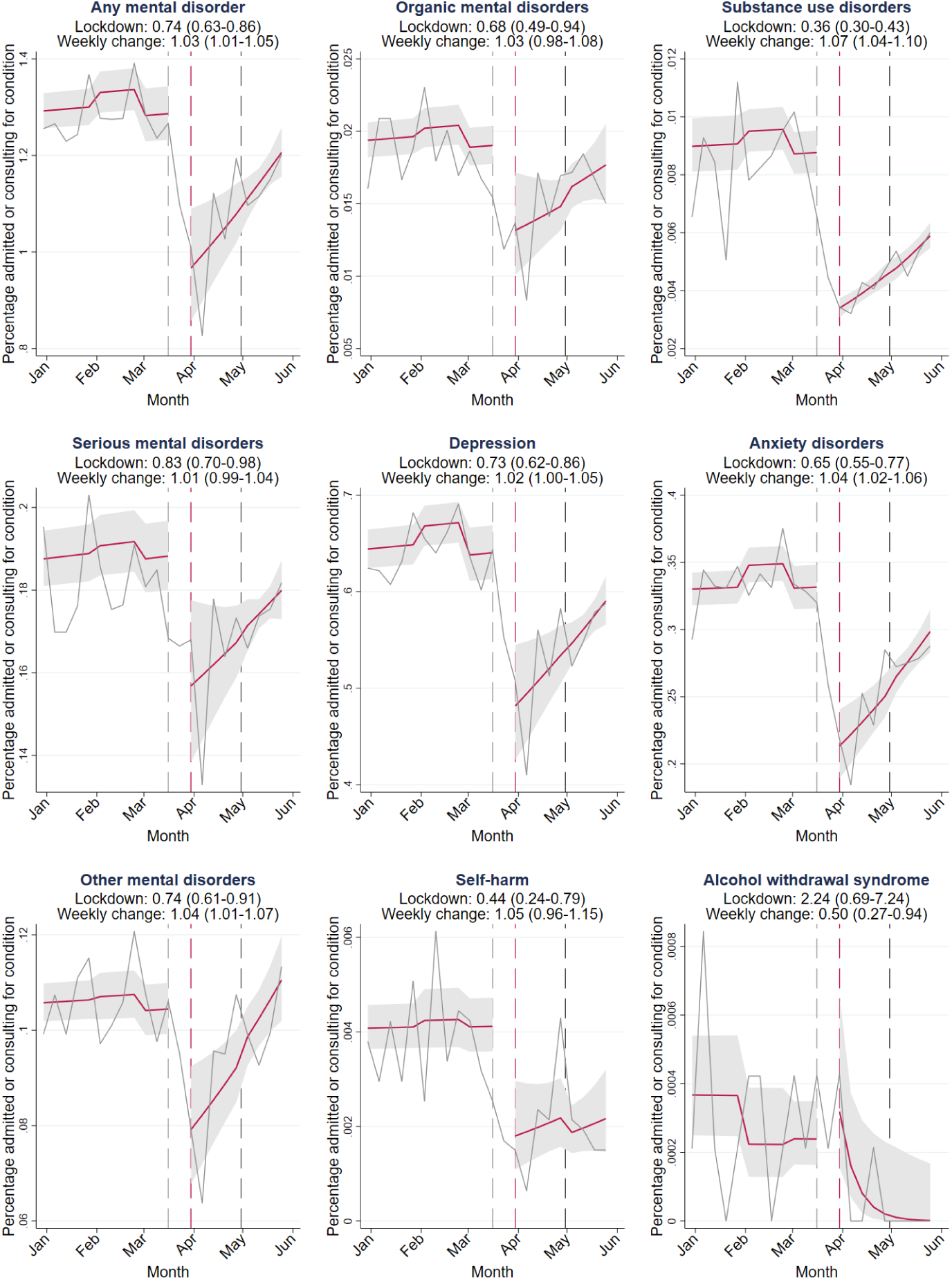
Interrupted time-series analysis for changes in mental health care use during the lockdown. Solid grey lines represent percentages of the study population admitted to a hospital or consulting outpatient care for condition in each week between January 1, 2020, and June 1, 2020. Red lines depict the estimated average percentage admitted or consulting outpatient care for the condition in a week with 95% CIs (grey shaded areas). Area between the dashed grey and dashed red line: data not used to account for anticipatory behaviour. Dashed red line: first Monday during lockdown (March 30, 2020). Dashed black line: beginning of lockdown level 4 (April 30, 2020). Lockdown: odds ratios (OR) for the immediate effect of the lockdown on hospital admission and outpatient consultation rates. Weekly change: OR for the weekly change in the odds of hospital admission and outpatient consultation rates during the lockdown (week 14-22 in 2020). 95% confidence intervals for ORs in parentheses.

Decreases in mental health care use for any condition were slightly higher in men than in women (supplementary Figure 1). The odds ratio for the effect of the lockdown on hospital admissions for any mental disorders was 0.39 (95% CI 0.30–0.51) in men and 0.44 (95% CI 0.37–0.52) in women. The lockdown effect on outpatient consultations was also slightly more pronounced in men (OR 0.77; 95% CI 0.65–0.91) than in women (OR 0.79; 95% 0.68–0.92).

The results from the Mansfield model were comparable to the results of our primary analysis (supplementary Table 1). There was no evidence for changes in mental health care utilisation in the same period in 2019 (weeks 14-22) as the lockdown was introduced in 2020 (supplementary Table 2).

Observed weekly hospital admissions rates, outpatient consultation rates, and overall mental health care utilisation rates for each of the conditions in each week between January 1, 2017, and June 28, 2020 are shown in the Appendix (supplementary Figures 2-4).

## Discussion

To the best of our knowledge, this is the first study examining the effects of the COVID-19 lockdown on mental health care utilisation in an African country. Hospital admission, outpatient consultation rates, and overall mental health care utilisation rates for mental disorders decreased after the introduction of the COVID-19 lockdown measures in South Africa in March 2020. The drop in rates was larger for hospital admissions than for outpatient consultations. We demonstrated that hospital admissions and outpatient care consultations for mental disorders dropped simultaneously, thereby excluding the possibility that either absorbed drops in the other. For most conditions, mental health care utilisation rates did not recover to pre-pandemic levels by June 1, 2020. Hospital admissions and outpatient consultations for alcohol withdrawal syndrome increased following the ban on alcohol sales in South Africa, but the statistical uncertainty around these estimates was too large to draw definite conclusions.

Our estimates of the magnitude of reductions in mental health care contacts during the lockdown are similar to estimates reported in other settings. A study from South Korea reported reductions in outpatient care visits for depression, anxiety disorders, and serious mental disorders close to our estimates of 15% to 30% reduction (Seo *et al*. 2021). A study from the UK reported a slightly higher reduction in psychiatric outpatient care visits from 20% to 46% (Mansfield *et al*. 2021). Our estimate of the decline in psychiatric hospital admissions of 58% corresponds to a Canadian study that reported a 56-60% decline in psychiatric emergency presentations in children and adolescents (Finkelstein *et al*. 2021). A study from Germany reported a lower reduction in psychiatric hospitalisations of 25% following the introduction of COVID-19 measures (Zielasek *et al*. 2021).

Substantial reductions in health care utilisation rates likely represent a large unmet need for mental health care. It is unlikely that lower rates of health care utilisation reflect a decrease in underlying disease prevalence, given the evidence that COVID-19 pandemic and lockdowns negatively impact mental health, with several countries reporting increased rates of mental illness and psychological distress during the pandemic (Brooks *et al*. 2020; Hossain *et al*. 2020; Iob *et al*. 2020; Santomauro *et al*. 2021; Xiong *et al*. 2020). The unmet need for mental health care may have long-term consequences for people with mental illness and their families. Untreated serious mental disorders, including psychotic disorders and bipolar disorder, can lead to legal, social, emotional, and financial problems or suicide (Altamura *et al*. 2010; Penttilä *et al*. 2014). Timely treatment of mental disorders with a high prevalence, including anxiety disorders and depression, is also important. Patients with depression receiving timely treatment have better clinical outcomes and a faster recovery, higher stability in life and better social functioning (Ghio *et al*. 2015). Untreated anxiety disorders tend to recur over time and increase in symptom severity (Craske & Stein 2016). Long-term consequences of untreated anxiety disorders may include social isolation, suicidality, substance abuse and physical comorbidity (Benatti *et al*. 2016; Craske & Stein 2016).

The less pronounced decreases in the rate of outpatient consultations compared to hospital admissions might be explained by the shift from in-person consultations to telemedicine. Before the lockdown, health insurances would only reimburse in-person consultations, but this regulation was revised during the lockdown (Kinoshita *et al*. 2020). According to a recently published study, in South Africa, 60% of patients used telepsychiatry, and 70% of psychiatrists practiced telepsychiatry as of May 2020 (Kinoshita *et al*. 2020). Nevertheless, the 25% reduction in outpatient consultations observed in this study still leaves a substantial void for many patients. On the upside, comparatively modest reductions in outpatient care compared to inpatient care could also signify that telepsychiatric services have the potential to ensure access to mental health care during lockdowns. Telepsychiatric outpatient care can also ensure access to prescription medicines, as doctors were also allowed to prescribe medication via telepsychiatric services (Kinoshita *et al*. 2020).

Our estimate of the unintended effect of the ban on alcohol sales on health care contacts for alcohol withdrawal syndrome is broadly consistent with a study from India showing a doubling in hospital presentations for the management of alcohol withdrawal syndrome following the ban on alcohol sales. Although the statistical uncertainty of our estimate is too large to draw definite conclusions, it is remarkable that the health care utilisation rates for alcohol withdrawal syndrome doubled while rates for all mental disorders dropped. This is especially noteworthy as we worked with private-sector data. Individuals who can afford private health insurance should also be more likely to have the financial means to stockpile alcohol at home or to secure their supplies by illegal means during the ban on alcohol.

Strengths of our study include the large sample size, longitudinal data with extended pre-pandemic follow-up, and the quasi-experimental study design. The use of a large national private-sector care database enabled us to study the effect of the lockdowns on health care utilisation for uncommon serious mental disorders. The long pre-pandemic follow-up allowed us to compare trends of the previous three years to 2020. Although we worked with observational data, the use of interrupted time series models allowed for a quasi-experimental design, taking full advantage of the longitudinal nature of the data, and allowing for adjustment for long-term temporal trends and seasonality in health care utilisation. Finally, our findings were robust in several sensitivity analyses.

Our results have to be considered in light of the following limitations. First, we could not study the recovery of service utilisation as restrictions were eased to lockdown level 3 in July 2020 because our study period ended in end-June 2020. Second, our study only included data from a private-sector medical aid scheme, and thus our findings are not necessarily applicable to the public sector. Third, we could not distinguish between in-person and virtual outpatient care consultations and therefore could not evaluate to what degree telemedicine compensated for drops in in-person outpatient care consultations. Fourth, since we used routine insurance claim data, we cannot exclude the possibility that changes to administrative procedures or reimbursement practices that may have occurred during the COVID-19 pandemic have influenced our results. Fifth, we had no information on the geographic location of health care providers or the residence of beneficiaries and could not examine regional differences in health care utilisation.

Further studies are needed to examine the underlying mechanisms that limited access to mental health care during the lockdown. Such mechanisms may include changes in the care-seeking behaviour of patients, transport-related and financial barriers, decreased psychiatric bed capacity to reduce the risk of in-hospital COVID-19 transmission, and other changes in service delivery possibly due to the reallocation of health care staff to care for COVID-19 patients. In addition, qualitative work is needed to understand how people living with mental illness and their primary care-takers coped without access to mental health care – whether they self-managed or sought support from social networks, traditional healers, or religious communities (Kola *et al*. 2021). Future studies should also evaluate the long-term consequences of delayed mental health treatment due to COVID-19 lockdowns on clinical outcomes. Finally, strategies to deliver essential mental health services during pandemics is a critical area for future research.

## Conclusions

In summary, reduced mental health care consultations rates during the COVID-19 lockdown likely reflect a substantial unmet need for mental health services in South Africa’s private sector, with potential long-term consequences for mental health patients and their families. Steps to ensure access and continuity of mental health services during future lockdowns should be considered.

## Data Availability

All data were obtained through the International epidemiology Database to Evaluate AIDS Southern Africa collaboration (IeDEA-SA, https://www.iedea-sa.org/). Data cannot be made available online because of legal and ethical restrictions. To request data, readers may contact IeDEA for consideration by filling out the online form available at https://www.iedea-sa.org/contact-us/
https://www.iedea-sa.org/

## Required Statements

## Acknowledgments

We thank Mansfield and colleagues (Mansfield *et al*. 2021) for making their statistical code publicly available.

## Author Contributions

AH and AW wrote the first draft of the study protocol. All authors contributed to the final version of the protocol. NM prepared the database. AH and AW performed statistical analysis. AW and AH wrote the first draft of the manuscript, which was revised by all authors. All authors approved the final version of the paper for submission.

## Conflicts of Interest

None.

## Financial Support

This study is supported by the US National Institutes of Health (the National Institute of Allergy and Infectious Diseases, the Eunice Kennedy Shriver National Institute of Child Health and Human Development, the National Cancer Institute, the National Institute of Mental Health, the National Institute on Drug Abuse, the National Heart, Lung, and Blood Institute, the National Institute on Alcohol Abuse and Alcoholism, the National Institute of Diabetes and Digestive and Kidney Diseases, and the Fogarty International Center) under award number U01AI069924 (Drs Egger and Davies). Dr Haas was supported by an Ambizione Fellowship (award number 193381) and Dr Egger by special project funding (award number 189498) from the Swiss National Science Foundation.

## Ethical Standards

The authors assert that all procedures contributing to this work comply with the ethical standards of the relevant national and institutional committees on human experimentation and with the Helsinki Declaration of 1975, as revised in 200.

## Availability of Data and Materials

All data were obtained from the IeDEA-SA. Data cannot be made available online because of legal and ethical restrictions. To request data, readers may contact IeDEA-SA for consideration by filling out the online form available at https://www.iedea-sa.org/contact-us/.

## Supplementary appendix

**Supplementary Figure 1:**
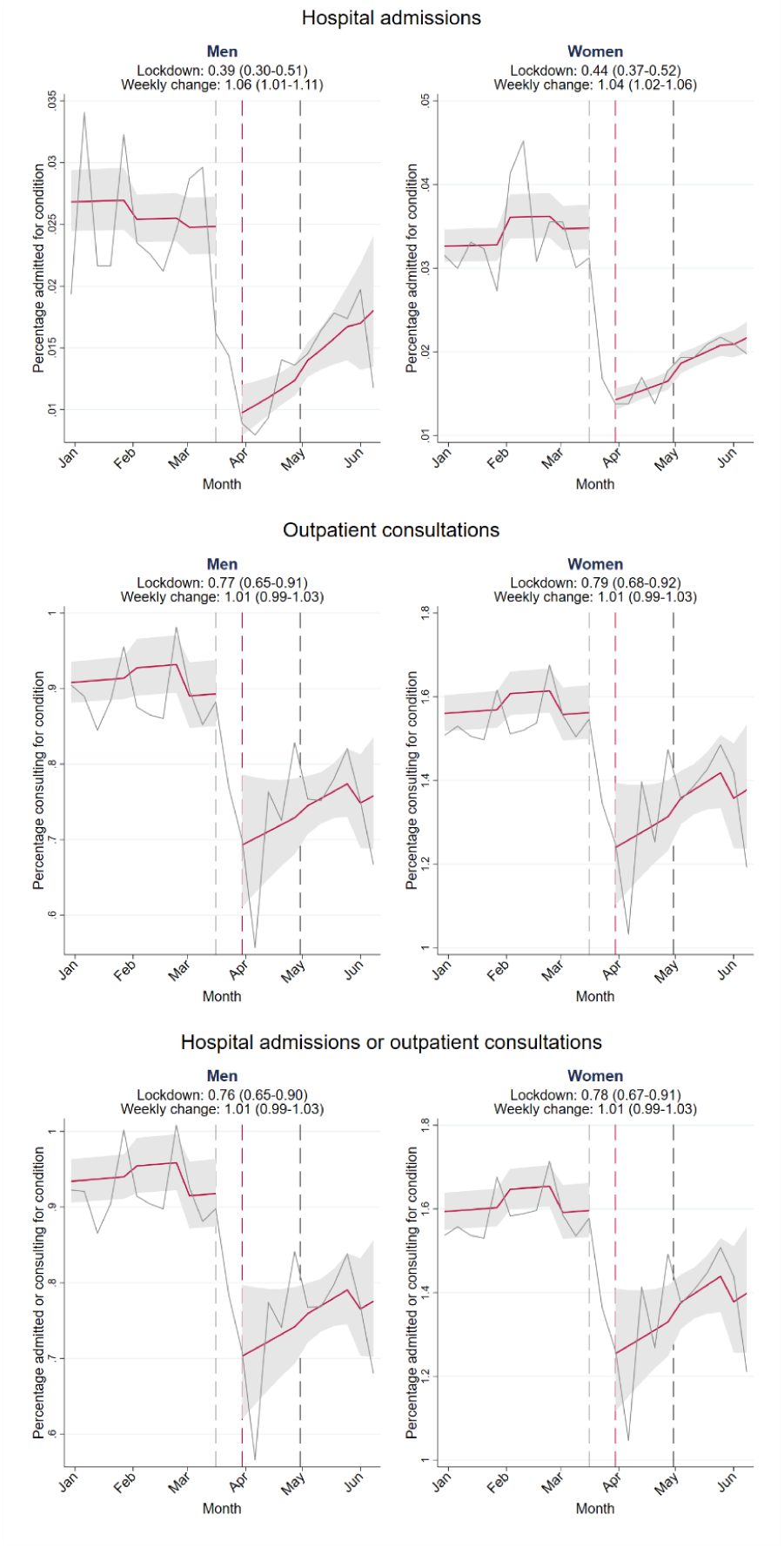
Interrupted time-series analysis for changes in mental health care use during the lockdown by sex. Solid grey lines represent percentages of the study population admitted to a hospital, consulting outpatient care, or admitted to a hospital or consulting outpatient care for any mental disorder in each week between January 1, 2020, and June 1, 2020. Red lines depict the estimated average percentage admitted to a hospital, consulting outpatient care, or admitted to a hospital or consulting outpatient care for any mental disorder in a week with 95% CIs (grey shaded areas). Area between the dashed grey and dashed red line: data not used to account for anticipatory behaviour. Dashed red line: first Monday during lockdown (March 30, 2020). Dashed black line: beginning of lockdown level 4 (April 30, 2020). Lockdown: odds ratios (OR) for the immediate effect of the lockdown on mental health care utilisation rates. Weekly change: OR for the weekly change in the odds utilizing mental health care during the lockdown (week 14-22 in 2020). 95% confidence intervals for ORs in parentheses.

**Supplementary Table 1.**
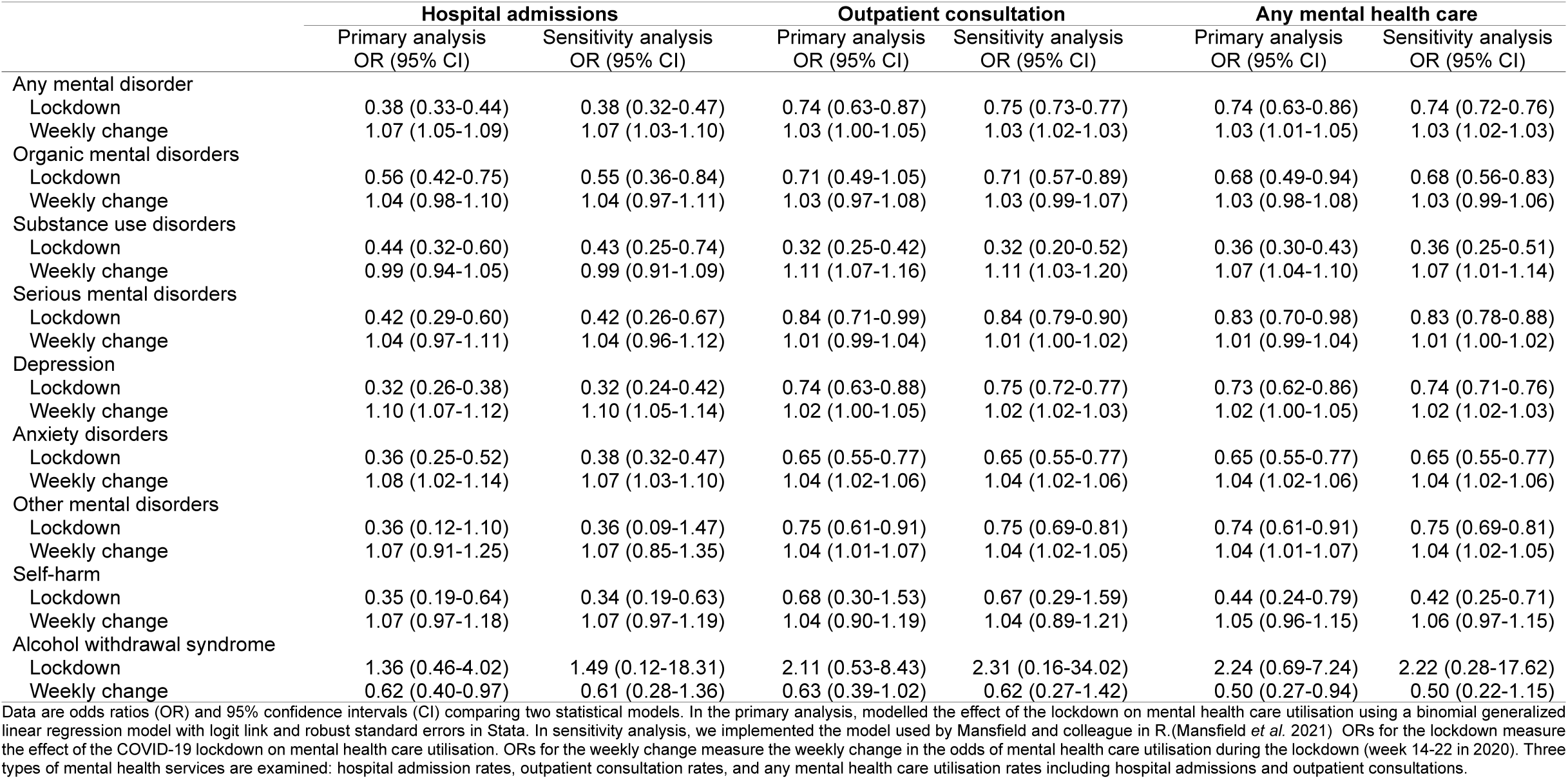
Comparison of statistical models of the effect of the COVID-19 lockdown on mental health care utilisation.

|  | Hospital admissions |  | Outpatient consultation |  | Any mental health care |  |
| --- | --- | --- | --- | --- | --- | --- |
|  | Primary analysis<br>OR (95% CI) | Sensitivity analysis<br>OR (95% CI) | Primary analysis<br>OR (95% CI) | Sensitivity analysis<br>OR (95% CI) | Primary analysis<br>OR (95% CI) | Sensitivity analysis<br>OR (95% CI) |
| Any mental disorder |  |  |  |  |  |  |
| Lockdown | 0.38 (0.33-0.44) | 0.38 (0.32-0.47) | 0.74 (0.63-0.87) | 0.75 (0.73-0.77) | 0.74 (0.63-0.86) | 0.74 (0.72-0.76) |
| Weekly change | 1.07 (1.05-1.09) | 1.07 (1.03-1.10) | 1.03 (1.00-1.05) | 1.03 (1.02-1.03) | 1.03 (1.01-1.05) | 1.03 (1.02-1.03) |
| Organic mental disorders |  |  |  |  |  |  |
| Lockdown | 0.56 (0.42-0.75) | 0.55 (0.36-0.84) | 0.71 (0.49-1.05) | 0.71 (0.57-0.89) | 0.68 (0.49-0.94) | 0.68 (0.56-0.83) |
| Weekly change | 1.04 (0.98-1.10) | 1.04 (0.97-1.11) | 1.03 (0.97-1.08) | 1.03 (0.99-1.07) | 1.03 (0.98-1.08) | 1.03 (0.99-1.06) |
| Substance use disorders |  |  |  |  |  |  |
| Lockdown | 0.44 (0.32-0.60) | 0.43 (0.25-0.74) | 0.32 (0.25-0.42) | 0.32 (0.20-0.52) | 0.36 (0.30-0.43) | 0.36 (0.25-0.51) |
| Weekly change | 0.99 (0.94-1.05) | 0.99 (0.91-1.09) | 1.11 (1.07-1.16) | 1.11 (1.03-1.20) | 1.07 (1.04-1.10) | 1.07 (1.01-1.14) |
| Serious mental disorders |  |  |  |  |  |  |
| Lockdown | 0.42 (0.29-0.60) | 0.42 (0.26-0.67) | 0.84 (0.71-0.99) | 0.84 (0.79-0.90) | 0.83 (0.70-0.98) | 0.83 (0.78-0.88) |
| Weekly change | 1.04 (0.97-1.11) | 1.04 (0.96-1.12) | 1.01 (0.99-1.04) | 1.01 (1.00-1.02) | 1.01 (0.99-1.04) | 1.01 (1.00-1.02) |
| Depression |  |  |  |  |  |  |
| Lockdown | 0.32 (0.26-0.38) | 0.32 (0.24-0.42) | 0.74 (0.63-0.88) | 0.75 (0.72-0.77) | 0.73 (0.62-0.86) | 0.74 (0.71-0.76) |
| Weekly change | 1.10 (1.07-1.12) | 1.10 (1.05-1.14) | 1.02 (1.00-1.05) | 1.02 (1.02-1.03) | 1.02 (1.00-1.05) | 1.02 (1.02-1.03) |
| Anxiety disorders |  |  |  |  |  |  |
| Lockdown | 0.36 (0.25-0.52) | 0.38 (0.32-0.47) | 0.65 (0.55-0.77) | 0.65 (0.55-0.77) | 0.65 (0.55-0.77) | 0.65 (0.55-0.77) |
| Weekly change | 1.08 (1.02-1.14) | 1.07 (1.03-1.10) | 1.04 (1.02-1.06) | 1.04 (1.02-1.06) | 1.04 (1.02-1.06) | 1.04 (1.02-1.06) |
| Other mental disorders |  |  |  |  |  |  |
| Lockdown | 0.36 (0.12-1.10) | 0.36 (0.09-1.47) | 0.75 (0.61-0.91) | 0.75 (0.69-0.81) | 0.74 (0.61-0.91) | 0.75 (0.69-0.81) |
| Weekly change | 1.07 (0.91-1.25) | 1.07 (0.85-1.35) | 1.04 (1.01-1.07) | 1.04 (1.02-1.05) | 1.04 (1.01-1.07) | 1.04 (1.02-1.05) |
| Self-harm |  |  |  |  |  |  |
| Lockdown | 0.35 (0.19-0.64) | 0.34 (0.19-0.63) | 0.68 (0.30-1.53) | 0.67 (0.29-1.59) | 0.44 (0.24-0.79) | 0.42 (0.25-0.71) |
| Weekly change | 1.07 (0.97-1.18) | 1.07 (0.97-1.19) | 1.04 (0.90-1.19) | 1.04 (0.89-1.21) | 1.05 (0.96-1.15) | 1.06 (0.97-1.15) |
| Alcohol withdrawal syndrome |  |  |  |  |  |  |
| Lockdown | 1.36 (0.46-4.02) | 1.49 (0.12-18.31) | 2.11 (0.53-8.43) | 2.31 (0.16-34.02) | 2.24 (0.69-7.24) | 2.22 (0.28-17.62) |
| Weekly change | 0.62 (0.40-0.97) | 0.61 (0.28-1.36) | 0.63 (0.39-1.02) | 0.62 (0.27-1.42) | 0.50 (0.27-0.94) | 0.50 (0.22-1.15) |

Data are odds ratios (OR) and 95% confidence intervals (CI) comparing two statistical models. In the primary analysis, modelled the effect of the lockdown on mental health care utilisation using a binomial generalized linear regression model with logit link and robust standard errors in Stata. In sensitivity analysis, we implemented the model used by Mansfield and colleague in R.(Mansfield *et al.* 2021) ORs for the lockdown measure the effect of the COVID-19 lockdown on mental health care utilisation. ORs for the weekly change measure the weekly change in the odds of mental health care utilisation during the lockdown (week 14-22 in 2020). Three types of mental health services are examined: hospital admission rates, outpatient consultation rates, and any mental health care utilisation rates including hospital admissions and outpatient consultations.

**Supplementary Table 2.** Interrupted time-series analysis for changes in mental health care utilisation in the same period in 2019 (weeks 14-22) as the lockdown was introduced in 2020.

|  | Hospital admissions<br>OR (95% CI) | Outpatient consultation<br>OR (95% CI) | Any mental health care<br>OR (95% CI) |
| --- | --- | --- | --- |
| Any mental disorder |  |  |  |
| Step change | 1.01 (0.77-1.33) | 1.02 (0.86-1.20) | 1.02 (0.86-1.20) |
| Weekly change | 1.00 (0.97-1.03) | 0.99 (0.97-1.01) | 0.99 (0.97-1.01) |
| Organic mental disorders |  |  |  |
| Step change | 0.97 (0.70-1.34) | 1.09 (0.88-1.35) | 1.05 (0.86-1.28) |
| Weekly change | 1.03 (0.99-1.08) | 0.98 (0.96-1.01) | 0.99 (0.97-1.02) |
| Substance use disorders |  |  |  |
| Step change | 1.17 (0.75-1.82) | 0.74 (0.51-1.06) | 0.95 (0.67-1.33) |
| Weekly change | 0.99 (0.93-1.05) | 1.05 (1.00-1.10) | 1.02 (0.97-1.06) |
| Serious mental disorders |  |  |  |
| Step change | 1.21 (0.91-1.62) | 0.97 (0.83-1.13) | 0.97 (0.84-1.13) |
| Weekly change | 0.99 (0.94-1.04) | 1.00 (0.98-1.01) | 1.00 (0.98-1.01) |
| Depression |  |  |  |
| Step change | 1.00 (0.74-1.35) | 1.02 (0.88-1.18) | 1.02 (0.87-1.18) |
| Weekly change | 1.00 (0.96-1.03) | 0.99 (0.98-1.01) | 0.99 (0.98-1.01) |
| Anxiety disorders |  |  |  |
| Step change | 0.97 (0.63-1.49) | 1.07 (0.86-1.32) | 1.07 (0.86-1.32) |
| Weekly change | 1.00 (0.94-1.06) | 0.99 (0.96-1.01) | 0.99 (0.96-1.01) |
| Other mental disorders |  |  |  |
| Step change | 1.89 (0.65-5.52) | 1.06 (0.86-1.31) | 1.06 (0.86-1.31) |
| Weekly change | 0.91 (0.79-1.04) | 0.99 (0.96-1.01) | 0.99 (0.96-1.01) |
| Self-harm |  |  |  |
| Step change | 1.28 (0.93-1.76) | 1.57 (0.80-3.10) | 1.19 (0.90-1.59) |
| Weekly change | 0.97 (0.93-1.01) | 0.98 (0.86-1.11) | 0.98 (0.94-1.02) |
| Alcohol withdrawal syndrome |  |  |  |
| Step change | 0.33 (0.04-2.60) | 2.62 (0.24-28.63) | 0.67 (0.16-2.93) |
| Weekly change | 1.02 (0.79-1.31) | 0.93 (0.64-1.36) | 0.97 (0.78-1.21) |

Data are odds ratios (OR) and 95% confidence intervals (CI) for changes in mental health care in the same period in 2019 (weeks 14-22) as the lockdown was introduced in 2020. ORs for the step-change measure the immediate change in mental health care use in week 14 in 2019. ORs for the weekly change measure the change in mental health care utilisation during weeks 14-22 in 2019. Three types of mental health services are examined: hospital admission rates, outpatient consultation rates, and any mental health care utilisation rates, including hospital admissions and outpatient consultations.

**Supplementary Figure 2:**
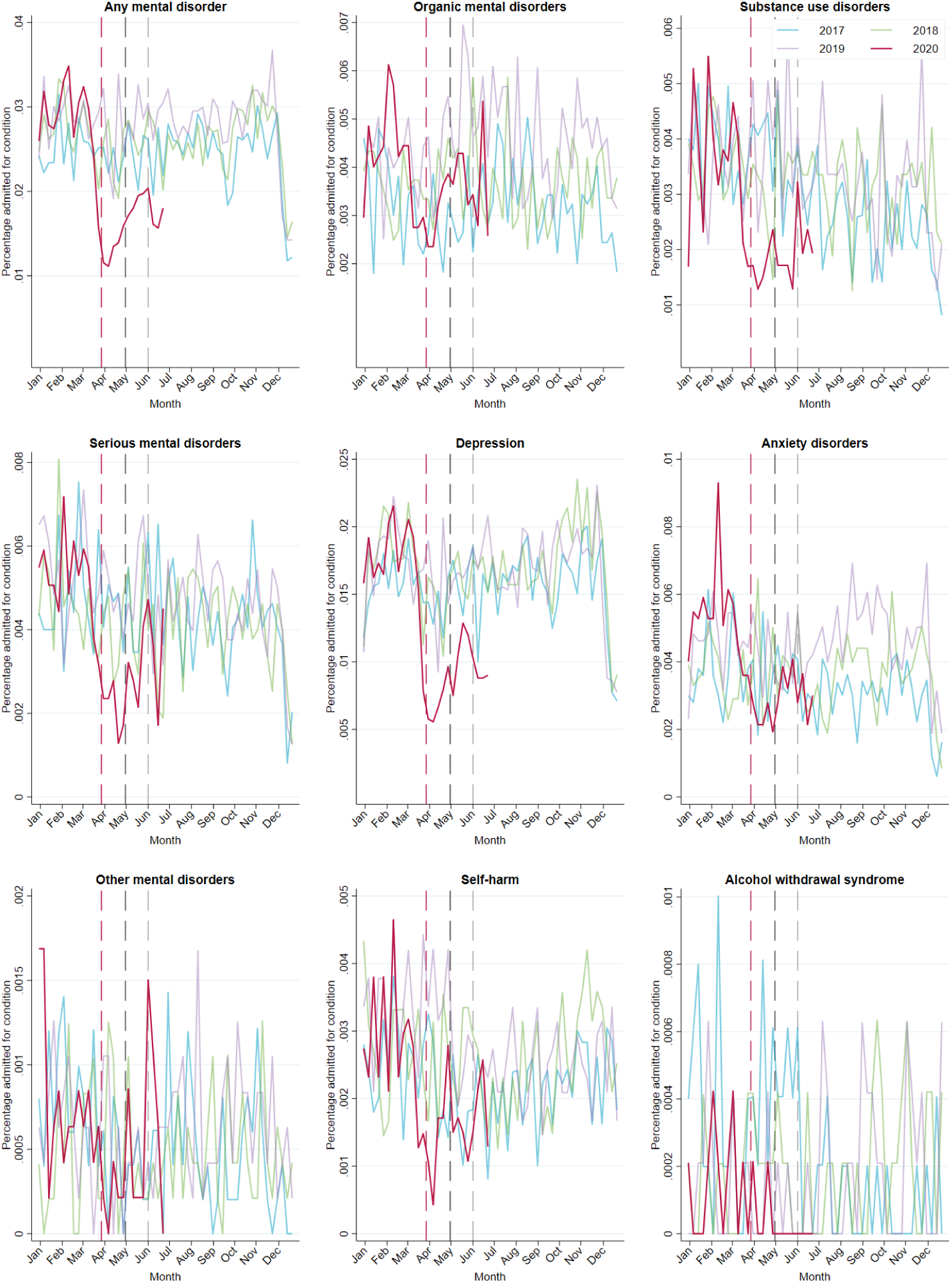
Hospital admission rates for selected mental health conditions. The coloured lines represent percentages of the study population admitted to a hospital for conditions in each week between January 1, 2017, and June 28, 2020. The dashed red, black, and grey lines show the beginning of lockdown levels 5, 4, and 3, respectively.

**Supplementary Figure 3:**
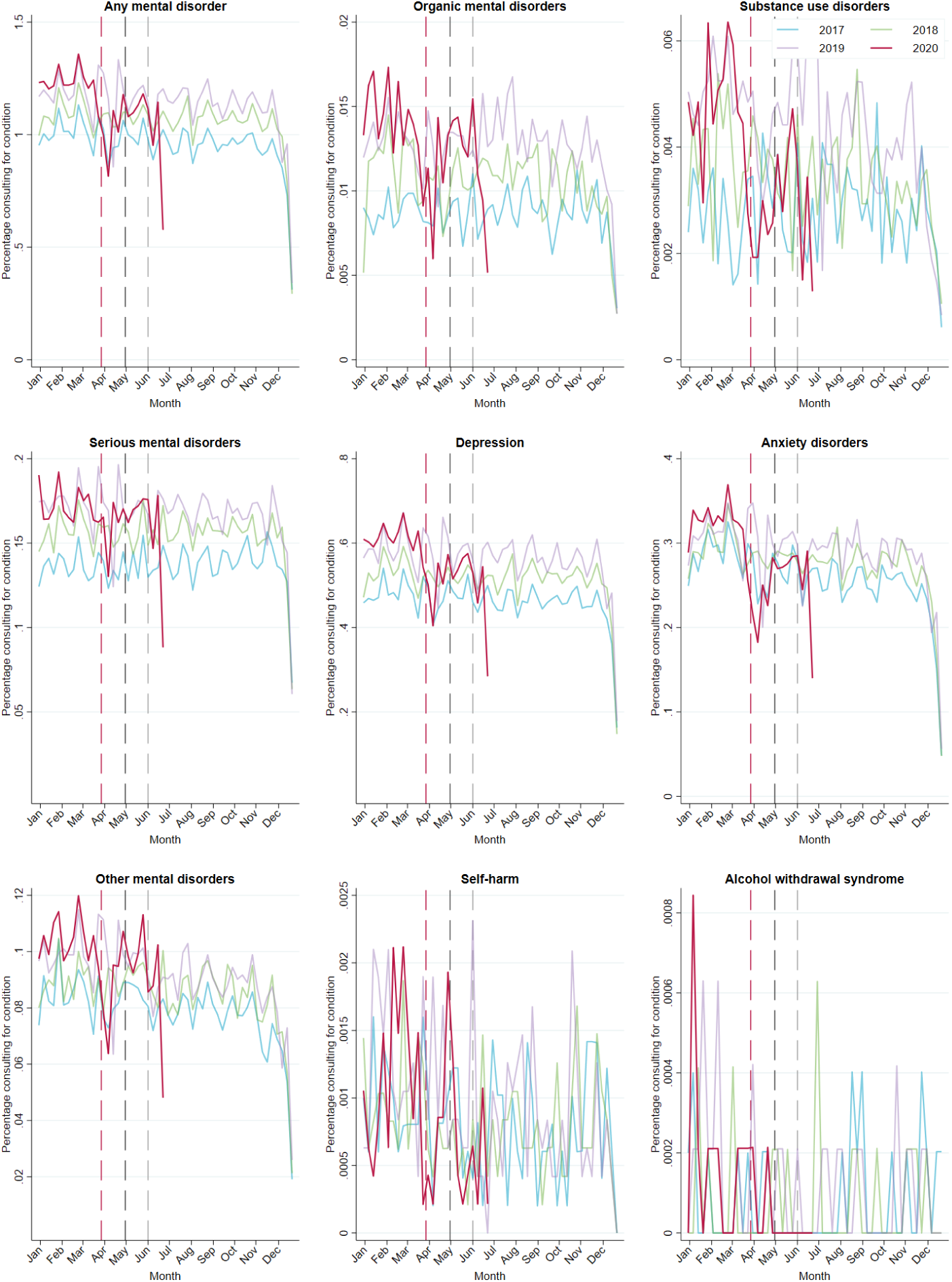
Outpatient consultation rates for selected mental health conditions. The coloured lines represent percentages of the study population consulting outpatient care for conditions in each week between January 1, 2017, and June 28, 2020. The dashed red, black, and grey lines show the beginning of lockdown levels 5, 4, and 3, respectively.

**Supplementary Figure 4:**
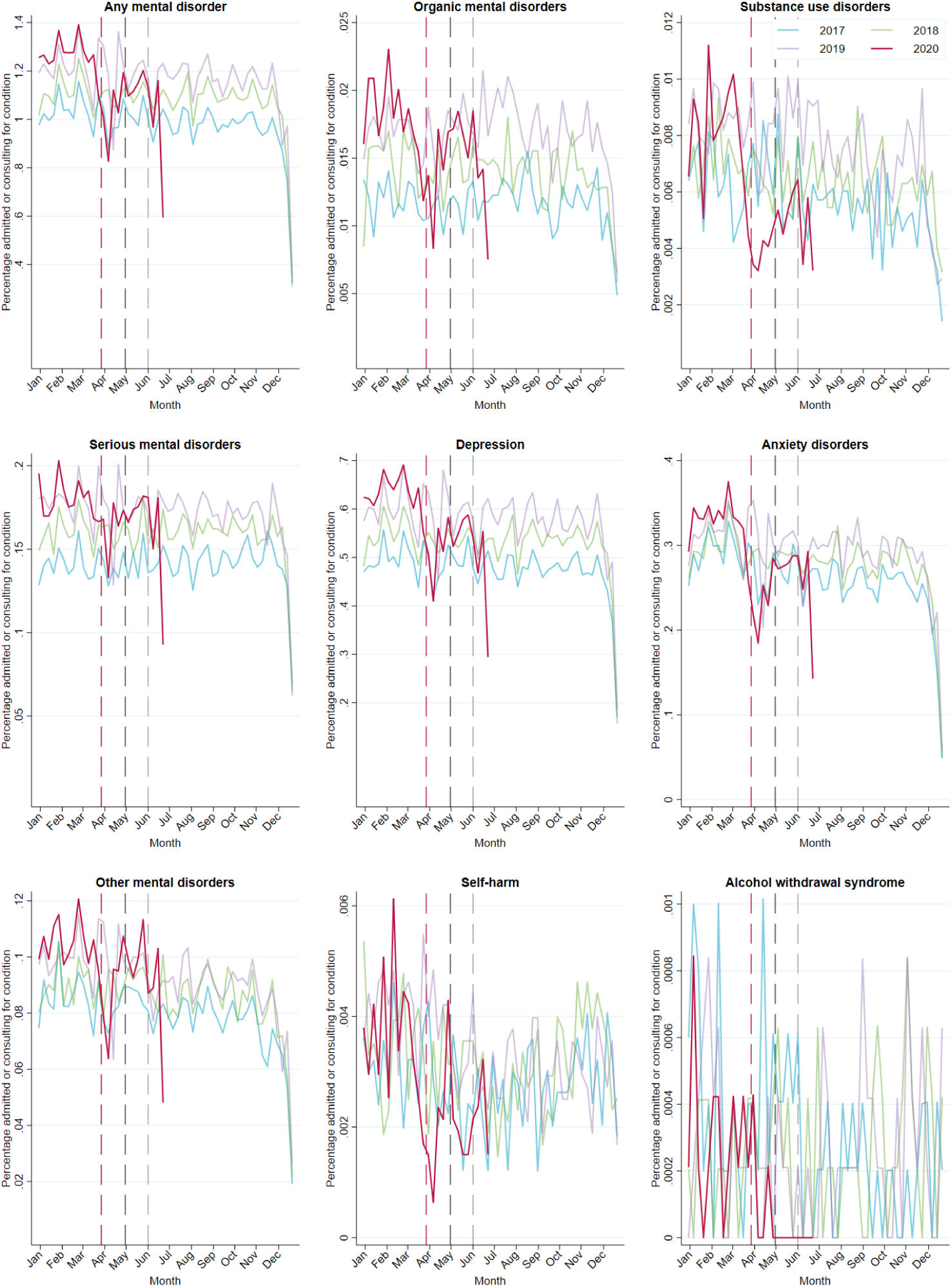
Health care utilisation rates for selected mental health conditions. The coloured lines represent percentages of the study population admitted to a hospital or consulting outpatient care for conditions in each week between January 1, 2017, and June 28, 2020. The dashed red, black, and grey lines show the beginning of lockdown levels 5, 4, and 3, respectively.

## Notes

### Competing Interest Statement

The authors have declared no competing interest.

### Author Declarations

The Human Research Ethics Committee of the University of Cape Town and the Cantonal Ethics Committee of the Canton of Bern granted permission to analyse the data

